# Time trends between vaccination coverage and voting patterns before and during the COVID-19 pandemic: analysis of COVID-19 and flu surveys in the United States

**DOI:** 10.1101/2022.09.05.22279620

**Authors:** Minttu M Rönn, Nicolas A Menzies, Joshua A Salomon

**Author notes:** **Corresponding author** Minttu Rönn, Department of Global Health and Population, Harvard T.H. Chan School of Public Health. The findings and conclusions in this report are those of the authors and do not necessarily represent the views of the funders or the authors’ affiliated institutions. **Ethics statement** The use of The US COVID-19 Trends and Impact Survey (CTIS) for analysis of COVID-19 symptoms and related behaviors was exempt from review by Harvard’s Institutional Review Board (protocol number IRB20-0592). Other data used in the study are publicly available aggregate estimates.

## Abstract

**Background:** We assessed the relationship between vaccination coverage and voting patterns: how has the association between COVID-19 vaccination and voting patterns changed during the pandemic, how does it compare to the association between flu vaccination coverage and voting patterns, and what can the time trends between flu vaccination and voting patterns tell us about the broader relationship between vaccination coverage and voting patterns.

**Methods:** We analyzed survey data on flu and COVID-19 vaccination coverage utilizing National Immunization Surveys for flu (NIS-FLU; years 2010-2021) and for COVID (NIS-ACM; 2021-2022), CDC surveillance of COVID-19 vaccination coverage (2021-2022) and US COVID-19 Trends and Impact Survey (CTIS; 2021-2022). We described the association between state-level COVID-19 and flu vaccination coverage and state-level voting patterns using Pearson correlation coefficient. We examined individual-level characteristics of people vaccinated for COVID-19 and for flu using logistic regression among responses in CTIS during April-June 2022. We analyzed flu vaccination coverage by age in NIS-FLU between 2010-2021, and its relationship with voting patterns to see whether there has been a departure from the secular pre-pandemic trend during the pandemic.

**Results:** Between May 2021 – June 2022 there was a strong and consistent correlation between state-level COVID-19 vaccination coverage and voting patterns for the Democratic party in the 2020 presidential elections. Pearson correlation coefficient was around 0.8 in NIS-ACM, CTIS and CDC surveillance with a range of 0.76–0.92. COVID-19 vaccination coverage in June 2022 was higher than flu vaccination coverage in all states and it had a stronger correlation with voting patterns (R=0.90 vs. R=0.60 in CTIS). There was a small reduction in the flu vaccination coverage between 2020-2021 and 2021-2022 flu seasons. In the individual-level logistic regression, vaccinated people were more likely to be living in a county where the majority voted for the Democratic candidate in 2020 elections both for COVID-19 (aOR .18, 95%CI 2.12-2.24) and for flu (aOR 1.38, 95%CI 1.36-1.41). We demonstrate a longstanding correlation between voting patterns and flu vaccination coverage. It varied by age with the strongest correlation in the youngest age groups. During the 2020-2021 flu season, all age groups, except for 5-12 years old, had a stronger correlation coefficient with voting patterns than in the previous years. However, the observed and predicted vaccination coverage show relatively modest differences in their correlation with vote share.

**Conclusions:** There are existing pre-pandemic patterns between vaccination coverage and voting patterns as demonstrated by the flu vaccination coverage for 2010-2021. During the pandemic COVID-19 vaccination has been more strongly correlated with vote share than the correlation observed for flu during and before the pandemic. The findings align with other research that has identified an association between adverse health outcomes and the political environment in the United States.

## INTRODUCTION

Politicization of attitudes towards COVID-19 vaccination is suggested to have contributed to geographical heterogeneities in vaccination coverage in the United States.^1^ Despite the US achieving one of the fastest initial rollouts of COVID-19 vaccination in early 2021,^2^ the country is lagging behind other high-income countries in coverage of two doses, and in coverage of additional vaccine doses.^3^ Jurisdictions with higher support for the Democratic party at the last presidential election have achieved a higher COVID-19 vaccination coverage,^4^ and survey results in November 2021 indicated that intention to receive a COVID-19 booster dose differed by party affiliation.^5^ Party voting patterns have also been associated with differences in mobility during the pandemic, and attitudes towards mitigation measures.^6–8^

While the COVID-19 pandemic and the vaccination program have created unique sociopolitical conditions and challenges, studies have identified an association between vaccination coverage and voting patterns before the pandemic. States where the Democratic candidate received the majority vote in the 2012 presidential election had a higher average vaccination coverage than states where the Republican candidate received the majority, for human papilloma virus vaccine (63.4% vs 56.0% for first dose in girls), meningococcal conjugate vaccine (90.1% vs 84.8%) and tetanus vaccine (79.3% vs 72.8%).^9^ Similarly, individuals who identified as Republican were less likely to report having received a flu shot compared to individuals who identified as Democrats between 2000-2012 (odds ratio 0.71, 95% confidence interval [95%CI] 0.61-0.84).^10^ During the H1N1 flu pandemic, acceptability of the H1N1 vaccine, and attitudes towards mass vaccination of Americans resulted in divisive discourse and party politics and media were seen to influence opinions.^11^

It is possible that the observed relationship between voting patterns and COVID-19 vaccination coverage simply reproduces pre-pandemic associations seen for other vaccines. Another limitation in disentangling these relationships is the cross-sectional nature of the earlier studies on vaccination coverage and voting patterns. Longer time trends remain under-explored, and such studies could explore the nature and evolution of the relationship between vaccination coverage and voting patterns. In this study, we aimed to answer three research questions: how has the relationship between COVID-19 vaccination and voting patterns changed during the pandemic, how does the association between COVID-19 vaccination coverage and voting patterns compare to association observed for flu vaccination coverage, what can the time trends between flu vaccination and voting patterns tell us about the broader relationship between vaccination coverage and voting patterns.

We evaluated how COVID-19 and flu vaccination coverage have changed before and during the pandemic in relation to presidential election vote share. We analyzed flu vaccination coverage over the past 10 flu seasons to examine whether the relationship between flu vaccination coverage and voting patterns has changed. The study can contribute to identifying emergent versus existing phenomena that contribute to low vaccination coverage in the United States.

## METHODS

### Data

#### Flu vaccination data

Annual influenza vaccination has been recommended for at-risk people since 1960, and populations recommended to receive the flu vaccine has evolved since.^12^ Currently annual flu vaccine is recommended for people over 6 months of age.^13^ We used two sources to estimate patterns of flu vaccination coverage (Supplementary Table S1):

i. National Immunization Surveys (NIS) are nationally representative telephone-based interviews. The component for influenza vaccination, NIS-FLU, has publicly available data with aggregate estimates of flu vaccine coverage over time for people 6 months and older by state, age and race/ethnicity.^14^ Public data are available from 2009 onwards, and in this study we used data from flu season 2010-2011 to 2020-2021 flu season. We omitted year 2009-2010 when two vaccines were available: the seasonal influenza vaccine and vaccine against the H1N1 strain, and the publicly available data for 2009-2010 flu season included people aged 50 and over only. We focused on vaccination coverage by age in our analyses given the small sample sizes for many of the race/ethnicity groups by state.
ii. The US COVID-19 Trends and Impact Survey (CTIS) is an online survey, implemented by the Delphi Group at Carnegie Mellon University, which recruited responses using the Facebook active user base of people aged 18 and older.^15^ The survey with individual-level responses includes a question about flu vaccination uptake during the 2020-2021 and 2021-2022 flu seasons. Uptake of flu vaccine for season 2020-2021 was in the survey for January 2021-August 2021, and a question about flu vaccine uptake for season 2021-2022 was in the survey for January-June 2022.

#### COVID-19 vaccination data

COVID-19 vaccines became available in December 2020, and vaccination is recommended for all people 6 months and older.^16^ Our definition of coverage was having received at least one dose of COVID-19 vaccine as this represents the most widespread estimate of achieved vaccination coverage, and provides a conservative estimate for our analyses. We used three data sources of COVID-19 vaccination coverage at state-level:

i. NIS Adult COVID-19 Module (NIS-ACM) estimates COVID-19 vaccine coverage, and public data are available for people aged 18 and older by state. Data are available from May 2021 to June 2022.^17^
ii. CTIS self-reported COVID-19 vaccination coverage, with individual-level responses received between January 2021 – June 2022 included in the analyses. Survey includes people aged 18 and older.^18^
iii. Aggregated data from the Centers for Disease Control and Prevention (CDC) on COVID-19 vaccination at state level. The data are considered the most complete dataset on COVID-19 vaccination coverage in the US. We used their total population coverage at state-level which is percent of population with at least one dose based on the jurisdiction where recipient lives.^15^

#### Voting patterns in presidential elections

To measure voting patterns, we used presidential election vote share at state- and county-level from a database maintained by the MIT Data Lab.^19^ We associated voting data from the most recent national election year. We considered the results from the most recent preceding election as the nearest indicator of political influence preceding each outcome, e.g. the 2020 presidential election vote share were used for 2020 and 2021 vaccination coverage data. Vote share was defined as percentage of votes for the Democratic candidate over total votes given in that geographic area.

#### Data availability

With the exception of CTIS microdata, the data are publicly available. Analytic code for the publicly available data are available in a GitHub repository.^20^

### Analyses

We conducted analyses in three stages to i) describe the overall patterns of correlation between COVID-19 and flu vaccination against voting patterns during the pandemic (2020-2022), ii) examine the similarities and differences in individual-level characteristics associated with vaccination status for COVID-19 and for flu in 2022, and iii) examine the longer time trend for flu vaccine coverage and voting patterns using flu surveys from 2010 onwards. All analyses were done in R.

#### 1. Descriptive analysis of correlation between vaccination coverage and presidential election vote share

We estimated the linear relationship between voting patterns and vaccination coverage. Surveys that rely on convenience sampling are less representative of the underlying population, as has been described for some of the largest COVID-19 vaccination surveys.^21^ To examine the consistency of relationship between COVID-19 vaccination coverage and vote share, we examined the strength and consistency of correlation across different datasets using the Pearson correlation coefficient. For COVID-19 vaccination coverage we calculated measures by month and for flu vaccination coverage we calculated measures by flu season. In CTIS data we additionally examined the cross-tabulation between flu and COVID-19 vaccination status at individual-level, categorizing people in one of the following states based on self-reported measures: not vaccinated for COVID-19 nor for flu (“none”), only received flu vaccine (“only flu”), only received COVID-19 vaccine (“only COVID-19”), or vaccinated against both COVID-19 and flu (“both vaccinations”). We present weighted estimates, to adjust for sampling and non-response in CTIS.

#### 2. Logistic regression of correlates with vaccination status in CTIS data

We performed two fixed-effects logistic regression analyses to examine individual-level factors associated with being vaccinated for COVID-19 and being vaccinated for flu. We included demographic variables that are expected to affect vaccination coverage (age, gender, race/ethnicity) and known socioeconomic correlates with vaccination and healthcare access (level of education, financial worry, employment status). We included voting patterns in the 2020 Presidential election at county-level: counties were categorized as those where the majority voted for the Republican candidate or for the Democratic candidate (≤50% and >50% of total votes in a county given to the Democratic candidate, respectively). We restricted the analysis to responses in CTIS survey wave 13 for April-June, 2022. The outcomes modeled were reporting having been vaccinated for COVID-19 (at least one dose, by the survey date), and having been vaccinated for flu since July 2021 (representing flu season 2021-2022); the models were run independently with the same explanatory variables. We used the R *survey* package to account for the survey design.^22^

#### 3. Analysis of flu vaccination coverage 2010-2021

We analyzed trends between flu vaccination coverage at state-level by age and vote share across flu seasons using Pearson correlation coefficient by flu season as an indicator of the strength of the relationship. We examined whether the relationships differed for the 2020-2021 flu season by predicting flu vaccination coverage for 2020-2021 season using data from the previous flu seasons and adjusting for the most recent presidential election vote share at state-level. The linear model had state-level flu vaccination coverage as the outcome, random intercept for states, random effects for age, and fixed effects for flu season, and state-level presidential vote share. Flu season was defined as a continuous variable from 2010.5 (season 2010-2011) to 2019.5 (season 2019-2020). We included presidential vote share of the previous presidential election as a continuous variable: 2008 presidential vote share for flu seasons until and inclusive of 2011-2012 flu season, 2012 presidential vote share until 2016-2017, 2016 presidential vote share until 2019-2020. Age was defined as a categorical variable. We excluded Washington DC from the analysis due the state being an outlier with >90% vote share for the Democratic party candidate. The model was used to predict the expected coverage at state-level by age for flu season 2020-2021 using 2020 presidential vote share. We compared the predicted and observed relationship between vaccination coverage and vote share by age using scatter plots and Pearson correlation coefficient.

## RESULTS

### 1. Correlation of state-level vaccination coverage with presidential 2020 vote share during the pandemic

COVID-19 vaccination coverage demonstrated a strong correlation with state-level 2020 vote share in CDC surveillance, NIS-ACM and CTIS (Figure 1). Between May 2021 – June 2022 the relationship remained relatively stable around 0.80 (range 0.76–0.92) in NIS-ACM and the CDC surveillance data, and around 0.90 (range 0.89-0.92) in CTIS. The observed relationship between vaccination coverage and vote share implied an average 8.6%, 5.8% and 4.7% increase in vaccination coverage for every 10% increase in Democratic vote share in data in June 2022 in CDC surveillance, NIS-ACM, and CTIS surveys, respectively.

**FIGURE 1.**
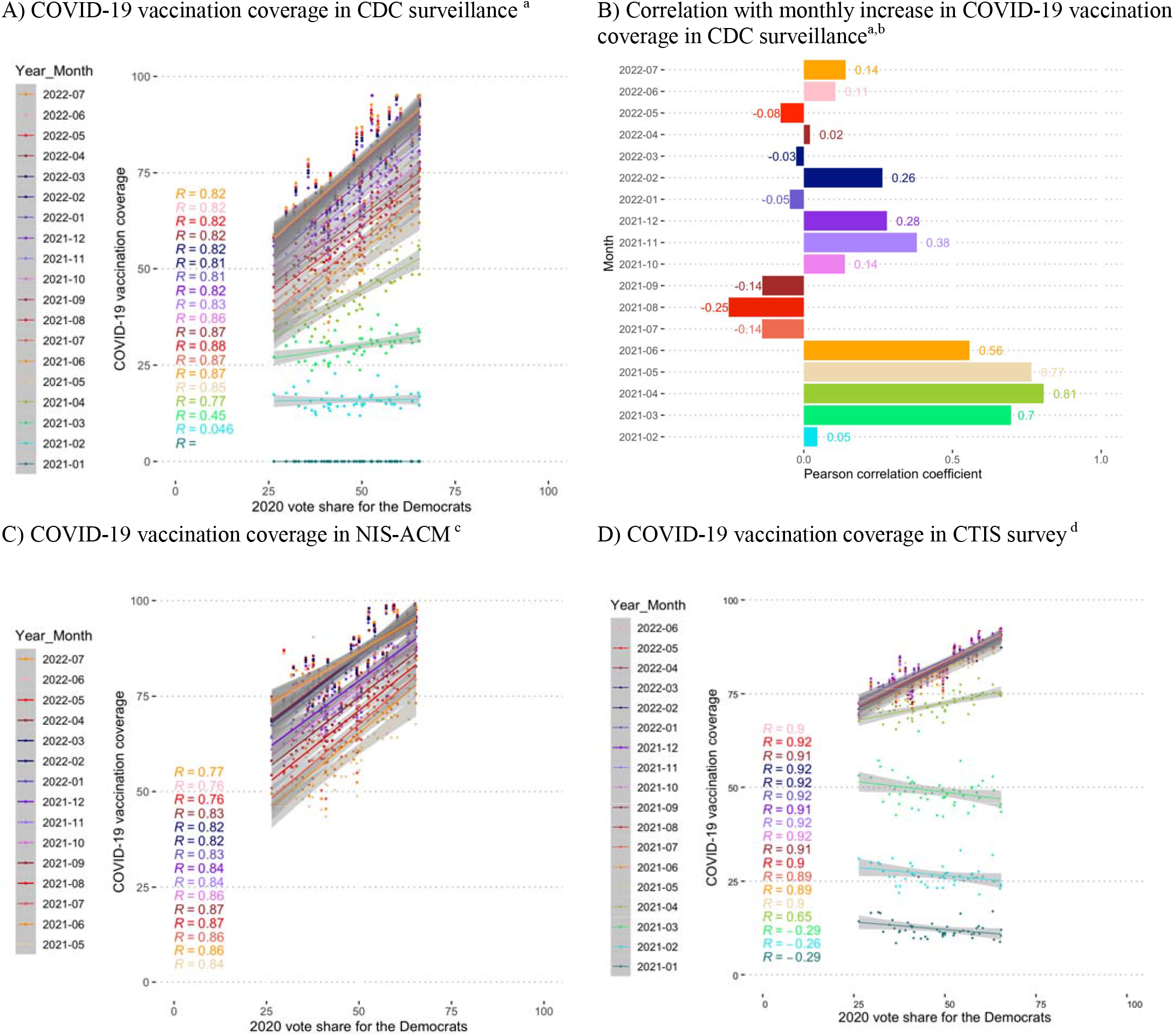
Comparison of correlation coefficient across state-level COVID-19 vaccination estimates by month. We have omitted Washington D.C. as an outlier. Pearson correlation coefficient (R) shown by month in panels A,C,D, and as a bar plot in panel B. Footnote: a) Percent of *total* population with at least 1 dose b) Absolute within state difference in COVID-19 vaccination coverage compared to the previous month in the CDC surveillance c) Received at least 1 dose, among people aged 18 and older d) Received at least 1 dose, among people aged 18 and older

While the relationship between vaccination coverage and Democratic vote share has been strong since early 2021, the relationship between increase in vaccine coverage and vote share were more variable over time in CDC surveillance data. The largest monthly increases in COVID-19 vaccination coverage took place between February-May 2021, and the vaccination coverage increased more in states with higher vote share for the Democratic party until June 2021 (Figure 1B, Supplemental Figure S1). However, from July to September 2021 the vaccination coverage increased more in states with lower vote share for the Democratic party; for example, ≥5% absolute increase in vaccination coverage was reported between July and August in eight states that had a majority vote for the Republican candidate in the 2020 presidential elections. Between October 2021 and February 2022, larger increases in coverage were in states with higher vote share for the Democratic party in all months except in January 2021. Since January 2021, vaccine eligibility expanded from the older age groups and at-risk groups to essential workers and to younger age groups at varied pace across the country, but all states offered COVID-19 vaccine to all adults by April 19, 2021.^23^ In October 29, 2021, Food and Drug Administration authorized emergency use of the Pfizer/BioNTech COVID-19 Vaccine for children aged 5-11 years old.^24^

We compared COVID-19 vaccination coverage to flu vaccination coverage among responses in CTIS in June 2022 (Figure 2A). COVID-19 vaccination coverage was higher in all states, and the correlation with the 2020 vote share was stronger for COVID-19 than for flu (R=0.90 vs. R=0.60). For flu vaccination, there was a 3.1% increase in flu vaccination coverage for every 10% increase in Democratic vote share. Most people who were vaccinated, had received both COVID-19 and flu vaccines, and the proportion of people with both vaccines was higher than what we would expect if uptake of the two vaccines was independent form each other (Supplement figure S2). Reporting not having received either of the vaccines was negatively correlated with the 2020 vote share (“None” in Figure 2B), and in the states with the lowest vote share for the Democratic candidate, over 20% of respondents reported not having received either COVID-19 nor flu vaccine. We also observed a small reduction in flu vaccination coverage from 2020-2021 to 2021-2022 flu season (Figure 2D). The reduction in flu vaccination coverage was larger in states with the lowest vote share for the Democratic party (Figure 2E)

**FIGURE 2.**
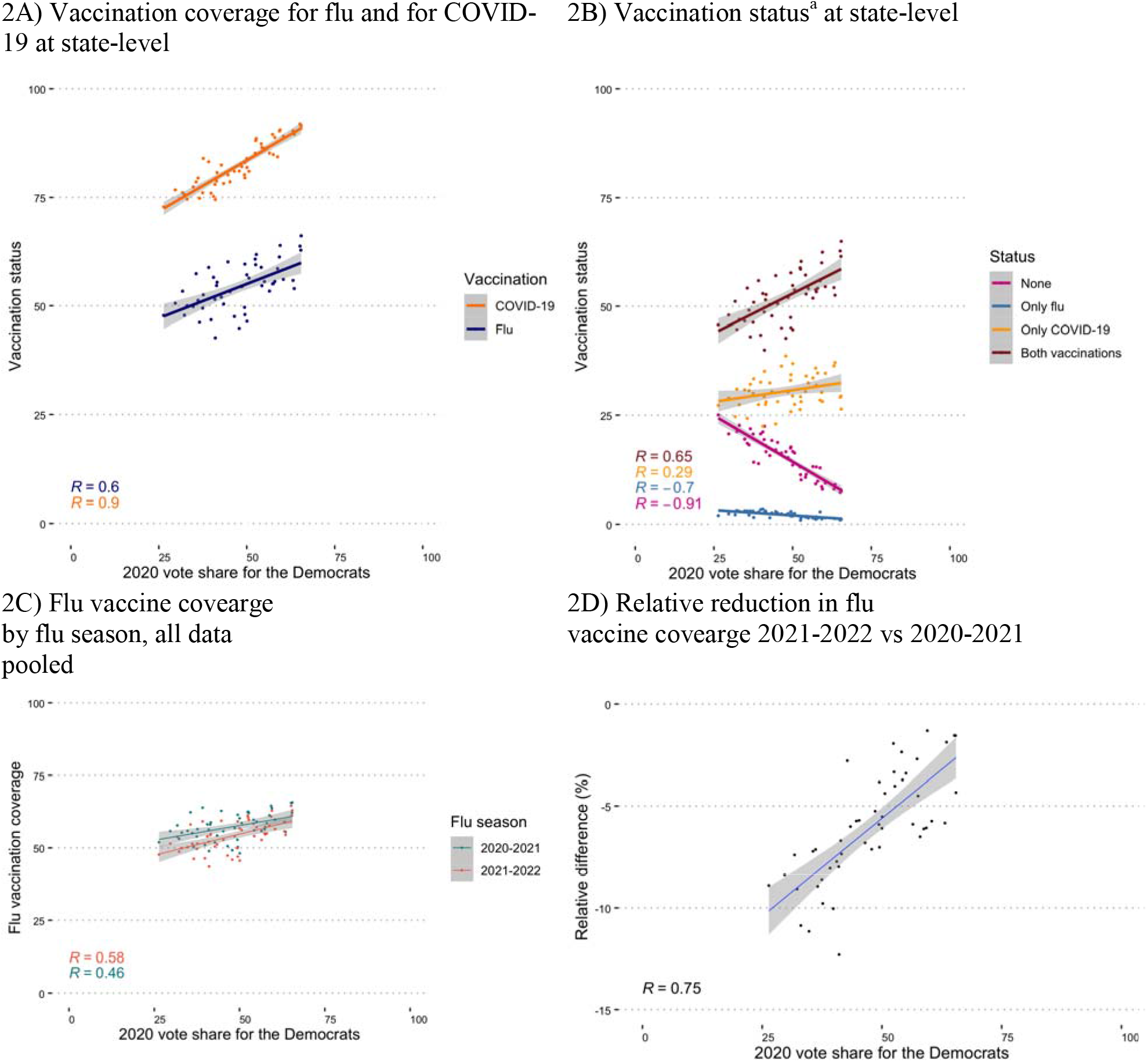
CTIS data vaccination coverage for flu and COVID-19 by state and state’s vote share for the Democratic party in 2020 presidential election. CTIS data for June 2022 in A-B, and aggregate responses for the flu seasons in C-D. Footnote: a) vaccination status defined as four discreet states: persons who reported receiving <u>neither</u> flu nor COVID-19 vaccine, <u>only</u> received the flu vaccines, only received the COVID-19 vaccine and those who received <u>both</u> vaccines.

### 2. Individual-level characteristics associated with vaccination coverage in 2022

Among CTIS responses during April-June 2022, the adjusted OR (aOR) of having received COVID-19 vaccination or flu vaccination (during flu season 2021-2022) is presented in Table 1. Increased age was associated for both COVID-19 and for flu, in similar fashion (Table 1). Men were less likely than women to be vaccinated for COVID-19, and were also less likely to be vaccinated for flu; aOR 0.58, (95%CI 0.56-0.59), and aOR 0.71 (0.70-0.72), respectively. People who reported being very worried about their household finances (aOR 0.71, 95%CI 0.69-0.73 and 0.72, 95%CI 0.70-0.73, compared to people who were somewhat or not worried over their household finances) were less likely to be vaccinated for COVID-19 and for flu, respectively. Similarly, people who had not completed high school (aOR 0.59, 95%CI 0.57-0.61 and 0.62, 95%CI 0.61-0.63, compared to people with high school or higher educational attainment) were less likely to be vaccinated for COVID-19 and for flu. Employment status in the past 4 weeks was not associated with either COVID-19 nor flu vaccination status.

**TABLE 1.** Adjusted odds ratios (aOR), and 95% CI from logistic regression models. Outcome is vaccination status for COVID-19 (A) or flu (B). Data of responses in wave 13, during April-June 2022 in CTIS. The models were additionally adjusted for the state of the respondent.

|  | **A) Vaccinated for COVID-19** |  |  | **B) Vaccinated for flu** |  |  |
| --- | --- | --- | --- | --- | --- | --- |
| **Sample size** | 473,078 |  |  | 474,665 |  |  |
|  | **aOR** | **95% CI** |  | **aOR** | **95% CI** |  |
| **Race/ethnicity** |  |  |  |  |  |  |
| White NH | 1.00 |  |  | 1.00 |  |  |
| Hispanic | 1.4 | 1.34 | 1.46 | 0.84 | 0.82 | 0.86 |
| Black NH | 1.21 | 1.15 | 1.29 | 0.69 | 0.67 | 0.72 |
| Multiple/Other NH | 0.47 | 0.44 | 0.49 | 0.63 | 0.6 | 0.66 |
| Asian NH | 4.28 | 3.77 | 4.87 | 1.31 | 1.24 | 1.39 |
| AI/AN | 0.65 | 0.58 | 0.72 | 0.71 | 0.65 | 0.77 |
| **Age group** |  |  |  |  |  |  |
| 18-24 | 1.00 |  |  | 1.00 |  |  |
| 25-34 | 1.05 | 0.99 | 1.12 | 1.17 | 1.11 | 1.23 |
| 35-44 | 1.22 | 1.15 | 1.29 | 1.44 | 1.36 | 1.51 |
| 45-54 | 1.52 | 1.43 | 1.61 | 1.65 | 1.57 | 1.74 |
| 55-64 | 2.16 | 2.04 | 2.29 | 2.41 | 2.3 | 2.54 |
| 65-74 | 3.62 | 3.4 | 3.86 | 4.3 | 4.09 | 4.53 |
| 75+ | 5.07 | 4.7 | 5.45 | 5.7 | 5.39 | 6.03 |
| **Sex** |  |  |  |  |  |  |
| Female | 1.00 |  |  | 1.00 |  |  |
| Male | 0.58 | 0.56 | 0.59 | 0.71 | 0.7 | 0.72 |
| **Very worried about finances for next month<sup>a</sup>** |  |  |  |  |  |  |
| No | 1.00 |  |  | 1.00 |  |  |
| Yes | 0.71 | 0.69 | 0.73 | 0.71 | 0.7 | 0.73 |
| **Education<sup>b</sup>** |  |  |  |  |  |  |
| HS or more | 1.00 |  |  | 1.00 |  |  |
| Less than HS | 0.59 | 0.57 | 0.61 | 0.62 | 0.61 | 0.63 |
| **In paid employment in the past 4 weeks<sup>c</sup>** |  |  |  |  |  |  |
| Yes | 1.00 |  |  | 1.00 |  |  |
| No | 1 | 0.98 | 1.03 | 1.1 | 1.08 | 1.12 |
| **Aggregate 2020 vote share at county-level<sup>d</sup>** |  |  |  |  |  |  |
| ≤50% for Democratic | 1.00 |  |  | 1.00 |  |  |
| >50% for Democratic | 2.18 | 2.12 | 2.24 | 1.38 | 1.36 | 1.41 |

- Survey question: How worried are you about your household's finances for the next month? Categorized as: Very worried (yes); Somewhat worried, not too worried, not worried at all (no) - Survey question: What is the highest degree or level of school you have completed? Categorized as: Less than high-school (HS) (yes); High-school graduate or equivalent and higher with multiple categories (no) - Survey question: In the past 4 weeks, did you do any kind of work for pay? Yes or No - 2020 Presidential election vote-share average at county level representing living in a Democratic (>50% vote share for Democrats) or Republican (≤50% vote share for Democrats) leaning county

Among CTIS responses, COVID-19 and flu vaccination patterns differed by race/ethnicity. Compared to non-Hispanic White respondents, COVID-19 vaccination was lower among multiple or other race/ethnicity (aOR 0.47, 95%CI 0.44-0.49) and American Indian/Alaska Native (aOR 0.65, 95%CI 0.58-0.72) respondents. Hispanic, Non-Hispanic Black and Non-Hispanic Asian respondents had a higher adjusted odds ratio for being vaccinated against COVID-19 compared to non-Hispanic White. For flu, compared to non-Hispanic White, vaccination was lower for most other race/ethnicity respondents, except for non-Hispanic Asian respondents (aOR 1.31, 95%CI 1.24-1.39).

Living in a county where the majority voted for the Democratic candidate in 2020 elections was more strongly associated with having been vaccinated against COVID-19 (aOR 2.18, 95%CI 2.12-2.24) than against flu (aOR 1.38 [1.36-1.41]), compared to living in a county where fewer than 50% of people voted for the Democratic candidate.

### 3. Analysis of state-level flu vaccination coverage 2010-2021

Long-term patterns in flu vaccination coverage based on NIS-FLU survey are displayed in Figure 3. Flu vaccination coverage and the relationship between coverage and voting patterns differed by age. People aged 50-64, and 65 and older had the highest flu vaccination coverage and the weakest correlation with state-level vote share (R <0.2 before 2018-2019, and R≥0.2 for 2019-2020 and 2020-2021). There was a stronger and relatively consistent correlation between flu vaccine coverage and voting patterns in younger age groups, compared to older ages, with a correlation coefficient of approximately 0.5 among age groups 17 years and younger. In 2010-2011 flu season, the strongest correlation coefficient was among people aged 5-12 years (R=0.54), and this remained similar in 2020-2021 (R=0.50). Since 2017-2018 flu season there has been an increase in flu vaccination coverage in the adult population, and this increase in coverage has been more pronounced in states with higher vote share for the Democratic party (Supplementary Figure S3 displays coverage by state and region).

**FIGURE 3.**
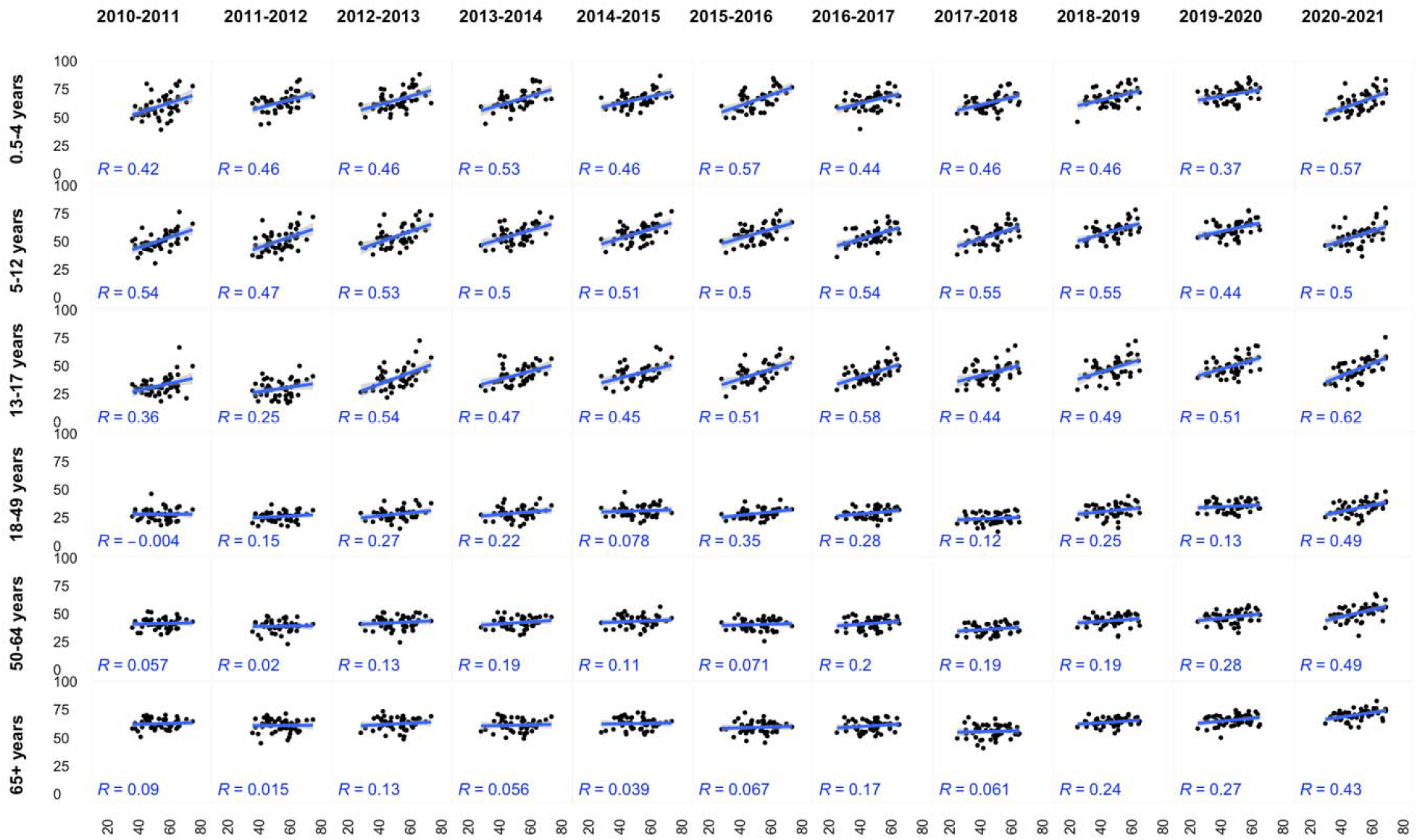
State-level correlation between 2020 vote share for Biden in 2020 presidential elections (x-axis, in %) and flu vaccine coverage (y-axis, in %) by age (rows) and by flu season (columns). Washington DC excluded from the figure.

**FIGURE 4.**
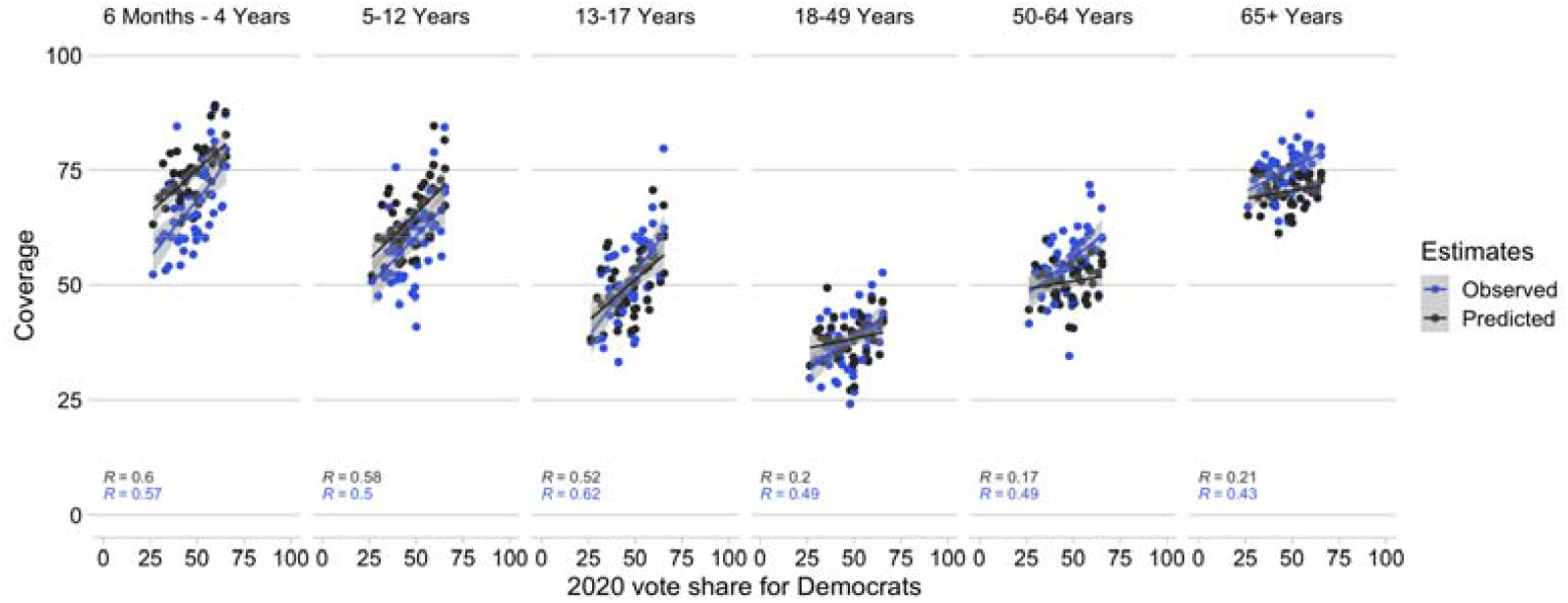
Predicted and observed flu vaccination coverage by age for the 2020-2021 flu season. Correlation between flu vaccine coverage and vote share for Democrats by age

During the 2020-2021 flu season, all age groups, except for 5-12 years old, had a stronger correlation coefficient with voting patterns than in the previous years. However, the observed and predicted vaccination coverage show relatively modest differences in their correlation with vote share (Figure 3). The largest divergence is for people aged 18 and over for whom the observed correlation between vaccination coverage and vote share is higher than the predicted correlation for 2020-2021 with the correlation coefficient in the predicted estimates being around 0.20 compared to the over 0.40 in the observed estimates.

## DISCUSSION

The relationship between COVID-19 vaccination coverage and voting patterns has stayed consistently strong throughout the pandemic. The states with higher vote share for the Democratic party candidate have seen larger monthly increases in the vaccination coverage, most prominently in the early phases of the vaccine rollout in early 2021 but also later when vaccination eligibility was expanded to younger age groups. When comparing state-level COVID-19 vaccination to flu vaccination during the pandemic, state-level COVID-19 vaccination coverage has approximately 50% higher correlation coefficient with the vote share compared to flu vaccination coverage. In June 2022, states with the least votes for the Democratic party candidate had significant gaps in vaccination coverage and this was apparent for both flu and COVID-19, and a high coverage of people who reported not having received either of the vaccines. In our analysis of individual-level data, living in Democratic leaning county remained more strongly associated with being vaccinated against COVID-19 than being vaccinated against flu when adjusting for individual-level demographic and socioeconomic variables.

We identified correlation between voting patterns and vaccination coverage for the entire duration of data, since 2010. For flu vaccination, the correlation varied by age with the strongest correlation in the youngest age groups. We did not identify a clear divergence in flu vaccination coverage in the first flu season of the pandemic (2020-2021) compared to the pre-pandemic flu seasons in NIS-FLU data. During 2020-2021, the relationship between vote share and flu vaccination coverage became stronger among the adult population, but it is possible this was part of a pre-pandemic trend of increasing vaccination coverage in the Democratic leaning states as opposed to increased vaccine uptake due to the pandemic.

Our analysis of CTIS data on changes in flu vaccination patterns during the pandemic showed a small reduction in vaccination coverage from the first pandemic flu season (2020-2021) to the second (2021-2022) with larger reduction in flu vaccination coverage in the states with lower vote share for the Democratic candidate. These data are limited due to their inability to identify reasons for the changes in vaccination coverage The flu vaccination coverage may also oscillate due to pandemic related temporary circumstances, and the relative recency of the COVID-19 pandemic makes identifying any emerging trends challenging. These findings highlight the need for further research to analyze whether flu vaccination coverage is changing.

The disparities for flu and COVID-19 are similar, and the individuals who are not vaccinated against either of the major respiratory diseases are more likely to live in the same geographies, and this can make the future COVID-19 and flu waves particularly problematic for these states. Measures of voting patterns are at ecological level, and reflect living in a state, or county, with an average vote share for either of the main party. Living in a Democratic or Republican leaning area is by default a proxy measure of distal factors. The findings of this study align with other research that has identified an association between adverse health outcomes and the political environment in the United States. Higher mortality rates persist in counties with lower vote share for the Democratic party, and this gap has grown during 2001-2019 as death rates have reduced less in Republican leaning counties than in Democratic leaning counties.^25^ The study identified diverse health outcomes such as heart disease, cancer, chronic lower respiratory tract diseases, unintentional injuries, and suicide as contributing to the difference in county-level mortality rates. Potential ways in which voting patterns may be associated with differences in health outcomes, and vaccination coverage, are via political decision-making, which influence health and social welfare policies via federal legislature and funding and at state-level via decisions about health care legislature, and budgeting, such as state-level expansion of Medicaid eligibility,^26^ and vaccination requirements by institution and employment differ by state.

In a global analysis, higher trust in the government was identified as having an inverse association with worse COVID-19 infection and mortality outcomes at country-level.^27^ Higher levels of interpersonal trust and government trust were also associated with higher COVID-19 vaccination coverage. United States has experienced worse health outcomes during the COVID-19 pandemic in comparison to many other high-income countries.^27^ For long-term management of COVID-19, understanding heterogeneities in COVID-19 vaccine coverage remains important. Despite different survey designs employed in the analyzed data, we observed a consistent association across data and for both COVID-19 and flu vaccines, and we demonstrated a pre-pandemic association with flu and voting patterns. Longer time series in the analyses can differentiate between aspects that were present pre-pandemic. This can help to disentangle potential true changes and account for the baseline levels of heterogeneity by voting patterns. It can also aid in monitoring vaccination coverage trends in the changing sociopolitical environment.

## Data Availability

With the exception of CTIS microdata, the data are publicly available. Analytic code for the publicly available data are available in a GitHub repository.

