## Supplemental Material for "Time trends between vaccination coverage and voting patterns before and during the COVID-19 pandemic: analysis of COVID-19 and flu surveys in the United States"

**Table S1.** Description of data surveys included in the analysis.

| Dataset | Vaccination measure | Population | Years | Age | Geography used | Race/ ethnicity | Outcome used | Survey methodology summary |
| --- | --- | --- | --- | --- | --- | --- | --- | --- |
| CDC Surveillance ^1^ | Covid-19 | Total population | 2020-2022  Ongoing | Total population | State | NA | End of month vaccine uptake coverage at state-level | Administrative Surveillance |
| NIS-ACM ^2^ | Covid-19 | General non-institutionalized population | 2021-2022  Ongoing | 18+ | State | 5 race/ethnicity groups | Monthly Vaccine uptake coverage at state-level  N.B. Monthly aggregate estimates provided are approximate, and do not necessarily represent a full calendar month (e.g. April 22 – May 29, 2011) | Phone interview |
| NIS-FLU ^3^ | FLU | General population | 2008-2021  Ongoing | 6 months+ | State | 5 race/ethnicity groups | Last vaccination coverage measure available of each flu season (May) at state-level | Phone interview |
| CTIS^4^ | Flu  Covid-19 | General population with Facebook account. | 2020-2022 | 18+ | County, State, | 6 race/ethnicity groups | Received any flu or any COVID-19 vaccination  County and State defined by the respondent’s FIPS | Survey administered on Facebook |

**FIGURE S1.** Absolute monthly difference in COVID-19 vaccination coverage, compared at state-level to previous month. Shading represents 2020 vote share for the Democrats. Data: CDC Surveillance

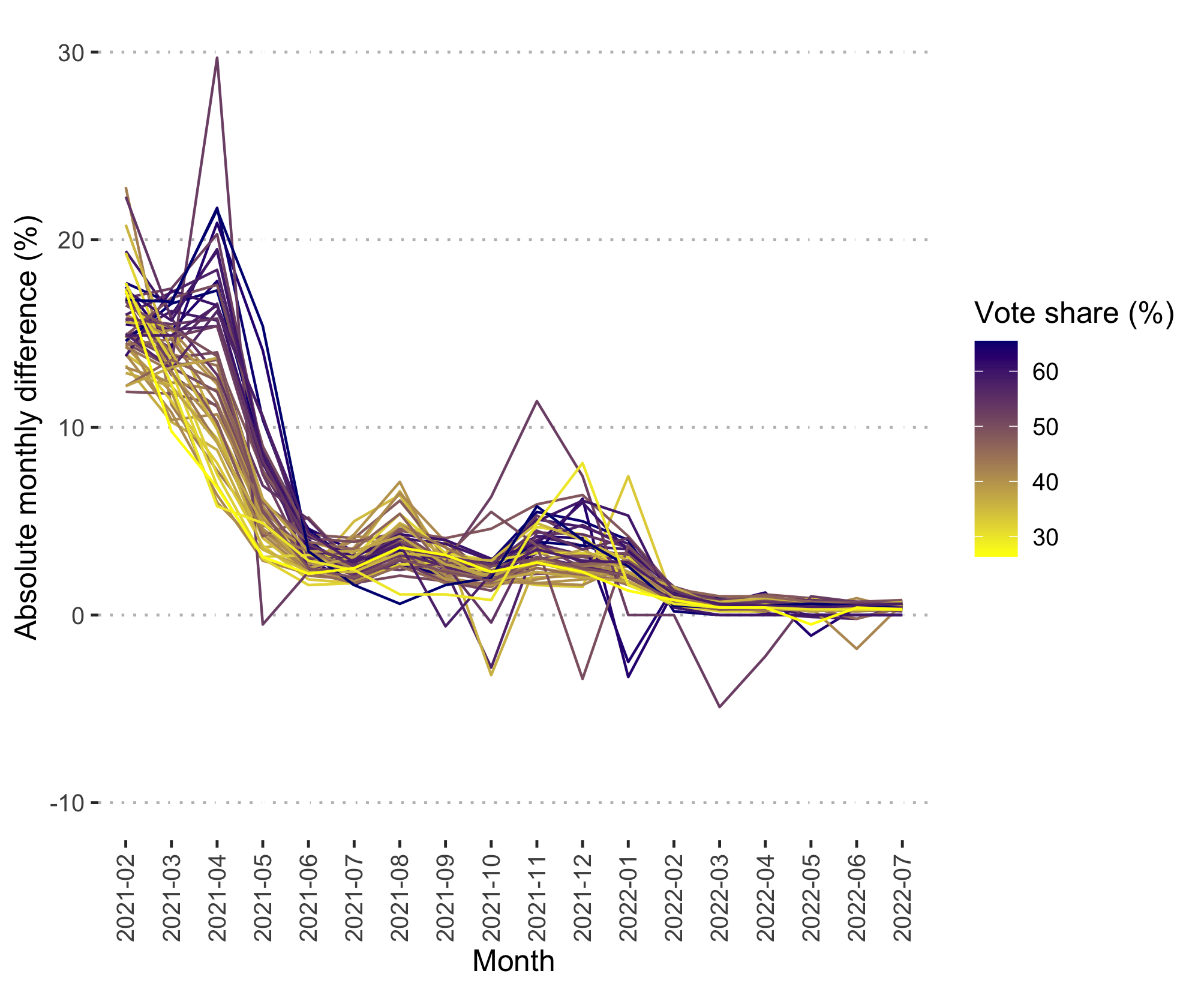

**Footnote**

The data are presented as they were reported, and no data cleaning were done to adjust for potential reporting changes and/or errors; e.g. reductions in population level vaccination coverage observed for some states.

**FIGURE S2.** CTIS data vaccination coverage for flu and COVID-19 by state and state’s vote share for the Democratic party in 2020 presidential election. CTIS data for June 2022.

Vaccination status at state-level: expected^a^ and observed compared to each other.

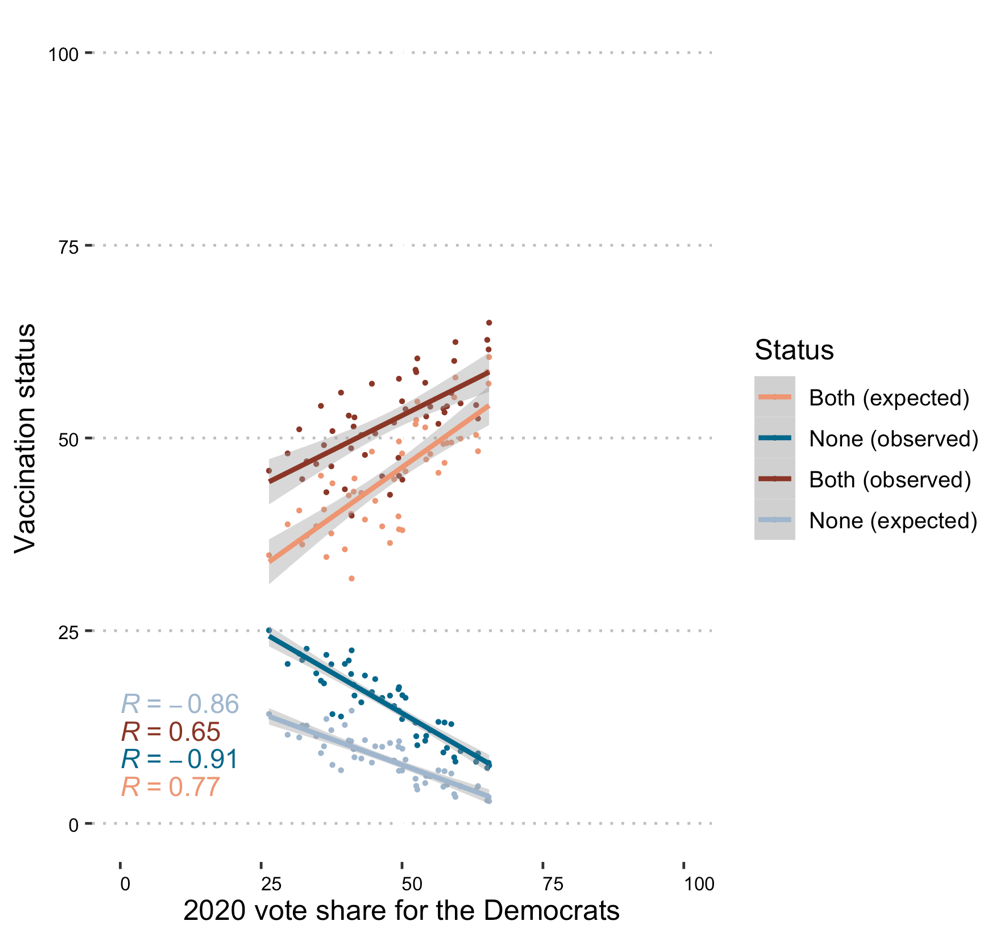

**Footnote**

a) Expected calculated based on individual vaccination coverage assuming independence: coverage of flu vaccine * coverage of COVID-19 vaccine.

Status defined as

Both = received both flu and COVID-19 vaccine

None = has not received flu nor the COVID-19 vaccine

**FIGURE S3.** State level flu vaccination coverage on y-axis by age (rows) and by HHS region (columns), organized by flu season 2010-2021 (x-axis). HHS regions* are ordered by 2020 vote share: Region 1 has the highest share for Democrats and Region 7 has the lowest vote share. The first flu season (2020-2021) during the COVID-19 pandemic is highlighted with orange. Data: NIS-FLU

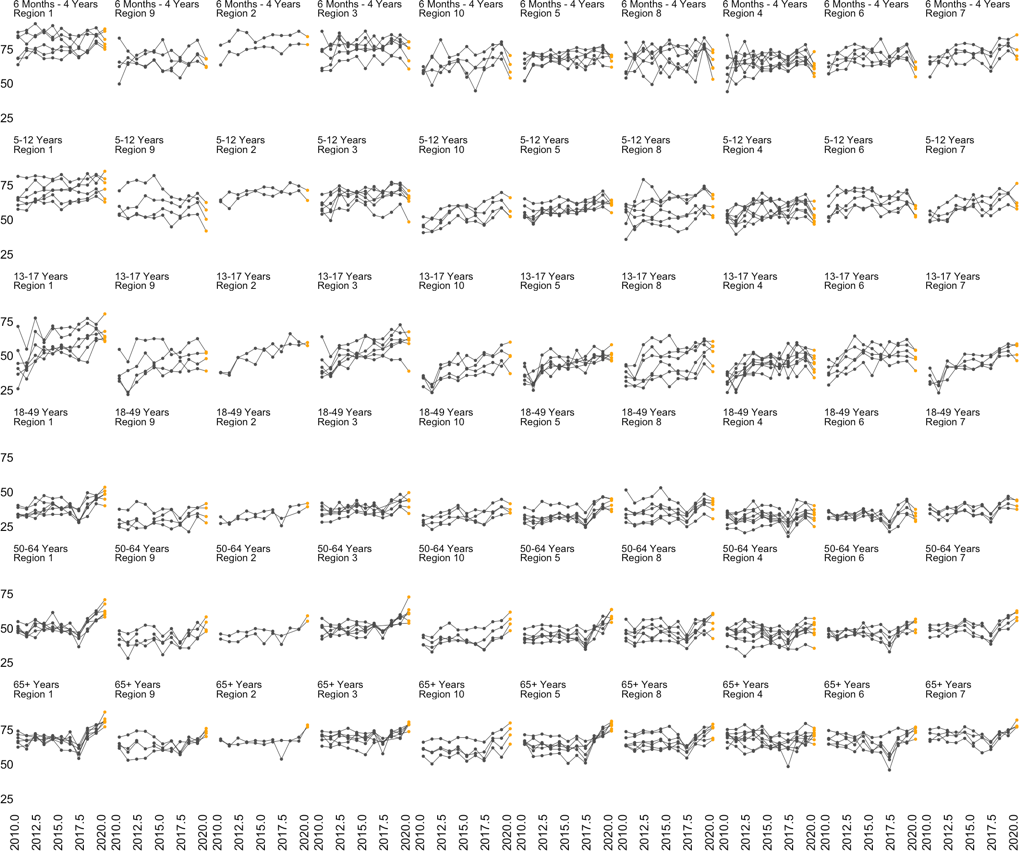

***) Footnote of HHS Regions**

Region 1: Connecticut, Maine, Massachusetts, New Hampshire, Rhode Island, and Vermont

Region 2: New Jersey, New York, Puerto Rico, and the Virgin Islands

Region 3: Delaware, District of Columbia, Maryland, Pennsylvania, Virginia, and West Virginia

Region 4: Alabama, Florida, Georgia, Kentucky, Mississippi, North Carolina, South Carolina, and Tennessee

Region 5: Illinois, Indiana, Michigan, Minnesota, Ohio, and Wisconsin

Region 6: Arkansas, Louisiana, New Mexico, Oklahoma, and Texas

Region 7: Iowa, Kansas, Missouri, and Nebraska

Region 8: Colorado, Montana, North Dakota, South Dakota, Utah, and Wyoming

Region 9: Arizona, California, Hawaii, Nevada, American Samoa, Commonwealth of the Northern Mariana Islands, Federated States of Micronesia, Guam, Marshall Islands, and Republic of Palau

Region 10: Alaska, Idaho, Oregon, and Washington
